# A booster dose is immunogenic and will be needed for older adults who have completed two doses vaccination with CoronaVac: a randomised, double-blind, placebo-controlled, phase 1/2 clinical trial

**DOI:** 10.1101/2021.08.03.21261544

**Authors:** Minjie Li, Juan Yang, Lin Wang, Qianhui Wu, Zhiwei Wu, Wen Zheng, Lei Wang, Wanying Lu, Xiaowei Deng, Cheng Peng, Bihua Han, Yuliang Zhao, Hongjie Yu, Weidong Yin

## Abstract

**Importance:** Whether herd immunity through mass vaccination is sufficient to curb SARS-CoV-2 transmission requires an understanding of the duration of vaccine-induced immunity, and the necessity and timing of booster doses. Objective: To evaluate immune persistence of two priming doses of CoronaVac, and immunogenicity and safety of a third dose in healthy adults ≥60 years. Design, setting, and participants: We conducted a vaccine booster study built on a single-center, randomized, double-blind phase 1/2 trial of the two-dose schedule of CoronaVac among healthy adults≥60 years in Hebei, China. We examined neutralizing antibody titres six months or more after the second dose in all participants. We provided a third dose to 303 participants recruited in phase 2 trial to assess their immune responses.

**Interventions:** Two formulations (3 μg, and 6 μg) were used in phase 1 trial, and an additional formulation of 1.5 μg was used in phase 2 trial. All participants were given two doses 28 days apart and followed up 6 months after the second dose. Participants in phase 2 received a third dose 8 months after the second dose.

**Main outcomes and measures:** Geometric mean titres (GMT) of neutralizing antibodies to live SARS-CoV-2 and adverse events were assessed at multiple time points following vaccination.

**Results:** Neutralizing antibody titres dropped below the seropositive cutoff of 8 at 6 months after the primary vaccination in all vaccine groups in the phase 1/2 trial. A third dose given 8 months or more after the second dose significantly increased neutralizing antibody levels. In the 3 μg group (the licensed formulation), GMT increased to 305 [95%CI 215.3-432.0] on day 7 following the third dose, an approximately 7-fold increase compared with the GMT 28 days after the second dose. All solicited adverse reactions reported within 28 days after a booster dose were of grade 1 or 2 severity.

**Conclusion and relevance:** Neutralizing antibody titres declined substantially six months after two doses of CoronaVac among older adults. A booster dose rapidly induces robust immune responses. This evidence could help policymakers determine the necessity and the timing of a booster dose for older adults.

**Trial registration:** ClinicalTrials.gov (NCT04383574).

## Introduction

Twenty COVID-19 vaccines (including inactivated, protein subunit, adenovirus vectored, and mRNA vaccines) are authorized globally, and widespread vaccination programmes are being implemented in over 200 countries/regions^1^. As of August 2, 4.18 billion doses of COVID-19 vaccines have been administered, with the highest coverage levels in Iceland (75% of the population fully vaccinated), United Arab Emirates (>70%), Israel (>60%), the United Kingdom and Spain (>50%), and the USA (nearly 50%)^2^. However, even with vaccination efforts in full force, a theoretical threshold for halting the spread of SARS-CoV-2 is uncertain and depends on factors such as how long vaccine-induced immunity lasts, and whether boosters are necessary^3^.

As vaccination campaigns gain speed, these pressing questions on vaccine-induced immune persistence and necessity of boosters become a focus of attention^4,5^. For the Moderna mRNA-1273 vaccine, vaccine-induced neutralizing antibody declined slightly but remained detectable in all participants 180 days after the second dose^6,7^. For Pfizer BNT162b2 and AstraZeneca ChAdOx1 nCoV-19 vaccines, antibody levels declined by 55% and 84% between 21–41 days and 70 days or more after the second dose^8^.

We previously reported results of our phase 2 clinical trial of CoronaVac, an inactivated vaccine against COVID-19 developed and produced by Sinovac Life Sciences, Beijing, China. That study showed that among 18–59-year-old subjects, neutralizing antibody titres declined to low levels 6 months after the second dose and a third dose was well tolerated and highly effective at recalling a SARS-CoV-2-specific immune response^9^. Here we report immune persistence of two priming doses of CoronaVac, and immunogenicity and safety of the third dose in healthy elderly adults aged 60 years or above, an age group at higher risk of severe illness and death from SARS-CoV-2 infection than young adults^10,11^.

## Methods

### Study design and participants

Our study is a vaccine booster study built on a single-center, randomized, double-blind phase 1/2 trial of CoronaVac which was initiated in Hebei, China on May 22, 2020, among healthy elderly adults aged 60 years or above. The design and main outcomes of the phase 1/2 trial have been published previously^12^. In summary, in phase I study, a dose escalation with two formulations, 3 μg (per 0.5 mL of aluminum hydroxide diluent) and 6 μg per dose, were assessed for the safety and tolerability. In phase 2, an additional formulation of 1.5 μg per dose was added together with two previous formulations, compared with placebo, to further assess the safety, tolerability, and immunogenicity. All participants in phase 1/2 trials were given two doses of CoronaVac 28 days apart.

Our vaccine booster study built on this phase 1/2 study comprised two parts. In the first part, we assess the immune persistence by examining the antibody titres at the 6-month visit after the second dose among participants recruited in the clinical trial phase 1/2. In the second part, we provided a third dose to participants recruited in the clinical trial phase 2 who completed 6-month follow up after the second dose.

Written informed consent was obtained from each participant before the third dose in the second part of the study. The clinical trial protocol and informed consent form were approved by the Ethics Committee of Hebei CDC (IRB2020–006).

### Randomization and masking

In phase 1, 72 participants were recruited and randomly allocated (1:1:1) to either a 3 μg, 6 μg, or placebo (aluminum hydroxide solution) group. In phase 2, 350 participants were randomly assigned (2:2:2:1) to either a 1.5 μg, 3 μg, 6 μg, or placebo group. Randomization codes for phases 1 and 2 were generated using block randomization and SAS software version 9.4; codes were assigned sequentially to participants. Study vaccine and placebo were identical in appearance. Details in the process of randomization and assignment were described previously^12^. All participants, investigators, and laboratory personnel were blind to study group assignment.

### Procedures

Vaccines (Vero-cell, inactivated CN02 strain of SARS-CoV-2, with an aluminum hydroxide adjuvant) of 1.5 μg, 3 μg, or 6 μg dosages or placebo were in prefilled syringes; appropriate vaccines or placebos were administered by intramuscular injection into deltoid muscles of subjects. Vaccination procedures for the first two doses have been described previously^12^. Additional exclusion criteria for the third dose are shown in Appendix 1.

Blood specimens were collected on day 0, 28, 56, and 208 post the first dose from participants in the study part 1 to evaluate 6-month immune persistence of the two-priming doses. Blood specimens were collected 7 or 14 days after the third dose among half of participants in the study part 2, and 28 days after a third dose among all participants in the study part 2. The objective of this timing was to evaluate rapidity of increase in neutralizing antibody titres after a third dose. The timing of each visit for assessment of immunogenicity and safety is shown in Appendix 2. Neutralizing antibody titres to live SARS-CoV-2 (virus strain SARS-CoV-2/human/CHN/CN1/2020, GenBank number MT407649.1) were quantified using a micro cytopathogenic effect assay, with laboratory assays previously described^12^.

The immunogenicity was assessed by the geometric mean titre (GMT) and seropositivity rate of neutralizing antibodies to live SARS-CoV-2 on day 180 after two-dose primary vaccination, and on days 7, 14, and 28 after the third dose. The positive cutoff titre for neutralizing antibodies to live SARS-CoV-2 was set to 1/8 in our study same as the value used in the previously published study^12^.

Safety of the third dose was assessed using the same method as for the first two doses^12^. For the first 7 days after the third dose, participants were required to record injection site adverse events (e.g., pain, swelling, or redness) and systemic adverse events (e.g., allergic reaction, cough, or fever) on diary cards. Between days 8 and 28, safety data were collected through spontaneous recording and reporting of adverse events by participants. Serious adverse events reported were collected until 6 months after the second dose for both phase 1 and 2 trials. Reported adverse events were graded according to China National Medical Products Administration guidelines^13^. Causal relations between adverse events and vaccination were assessed by the investigators.

### Outcomes

Results of safety and immunogenicity until 28 days after the first two doses have been published^12^. The immunogenicity endpoint was the geometric mean titre (GMT) and seropositivity rate of neutralizing antibodies to live SARS-CoV-2 on day 180 after two-dose primary vaccination, and on days 7, 14, and 28 after the third dose. The positive cutoff titre for neutralizing antibodies to live SARS-CoV-2 was 1/8.

The safety endpoints included any vaccine-related adverse event (adverse reaction) within 28 days after administration of the third dose of vaccine or placebo in phase 2 trial, and serious adverse events from immunization till 6 months after two doses in both phase 1&2 trials.

### Statistical analysis

We assessed immunogenicity endpoints in the per-protocol population (descriptions in Appendix 3), which included all participants who completed their assigned vaccinations, had antibody results available, and did not violate the trial protocol. In phase 1, we assessed immunogenicity 6 months after the second dose in an immune persistence set. In phase 2, the immunogenicity before administration of the third dose was assessed in a full analysis set, which included all participants who received the third dose. Immunogenicity of the third dose was assessed in the per-protocol population according to the protocol of each visit. We assessed serious safety events for 6 months after two doses in a safety population data set that included all participants who received at least one dose of vaccine or placebo. Safety assessment for the third dose was performed in a safety population data set of all subjects who received a third dose.

As stated in previous publication^12^, we did not determine the sample sizes on the basis of a statistical power calculation. Instead, we followed the requirements of the China National Medical Products Administration and Chinese Technical Guidelines for Clinical Trials of Vaccines—that is, recruitment of at least 20–30 participants in phase 1, and 300 participants in phase 2.

We used the Pearson χ^2^ test or Fisher’s exact test for analyses of categorical outcomes. We calculated 95% CIs for categorical outcomes using the Clopper-Pearson method. We calculated GMTs and corresponding 95% CIs using standard normal distributions of log-transformations of antibody titres. We used ANOVA to compare log-transformed antibody titres. T-test comparisons between groups were performed when comparisons among all groups showed any significant difference. Hypothesis testing was two-sided, and we considered p values less than 0.05 to be significant. We used R version 4.0.2 for all analyses.

An independent data monitoring committee consisting of one independent statistician, one clinician, and one epidemiologist was established before commencement of the study. Safety data were assessed and reviewed by the committee as the study proceeded.

## Results

### Participants

Between May 22, 2020 to April 22, 2021, 68 out of 72 participants being randomized finished the 6-month follow up and included in our part 1 immune persistence analysis with 21 in the 3 μg group, 23 in the 6 μg group, and 24 in the placebo group. In the part 2 of our study, 303 out of 350 participants being randomized received the third dose at 8 months or more after the second dose, with 85 in the 1.5 μg group, 90 in the 3 μg, 81 in the 3 μg group, and 47 in the placebo group (Figure 1). The mean age of participants received a third dose was approximately 67 years and characteristics of participants were similar between groups as described in Table 1.

**Table 1.** Baseline demographic characteristics for individuals who received the third dose in the phase 2 trial.

|  | 1.5 µg group<br>(n=85) | 3 µg group<br>(n=90) | 6 µg group<br>(n=81) | Placebo group<br>(n=47) | *P* value |
| --- | --- | --- | --- | --- | --- |
| Age, years |  |  |  |  | 0.735 |
| Mean (SD) | 66.3 (4.4) | 66.4 (4.4) | 66.3 (4.4) | 67.1 (4.7) |  |
| 60-64 | 33 (39%) | 35 (39%) | 31 (38%) | 16 (34%) |  |
| 65-69 | 32 (38%) | 29 (32%) | 33 (41%) | 15 (32%) |  |
| ≥70* | 20 (24%) | 26 (29%) | 17 (21%) | 16 (34%) |  |
| Sex |  |  |  |  | 0.633 |
| Male | 41 (48%) | 44 (49%) | 37 (46%) | 27 (57%) |  |
| Female | 44 (52%) | 46 (51%) | 44 (54%) | 20 (43%) |  |
| BMI, kg/m<sup>2</sup> |  |  |  |  | 0.341 |
| Mean (SD) | 25.6 (3.3) | 25.8 (3.6) | 25.5 (3.2) | 24.7 (3.4) |  |

\*The oldest participants were 80 years old, including one subject in the 6 µg group, and two subjects in the placebo group.

**Table 2.**
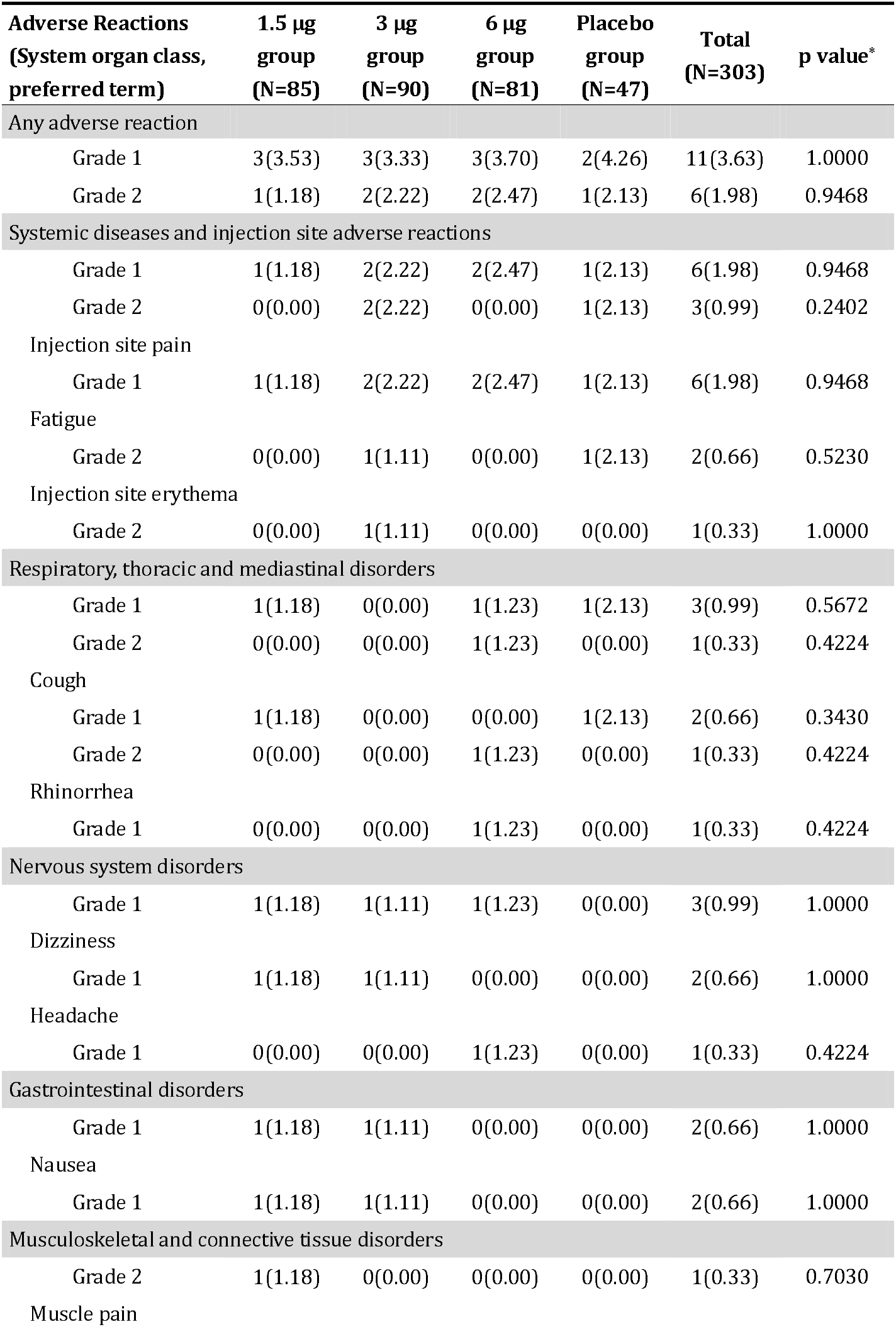
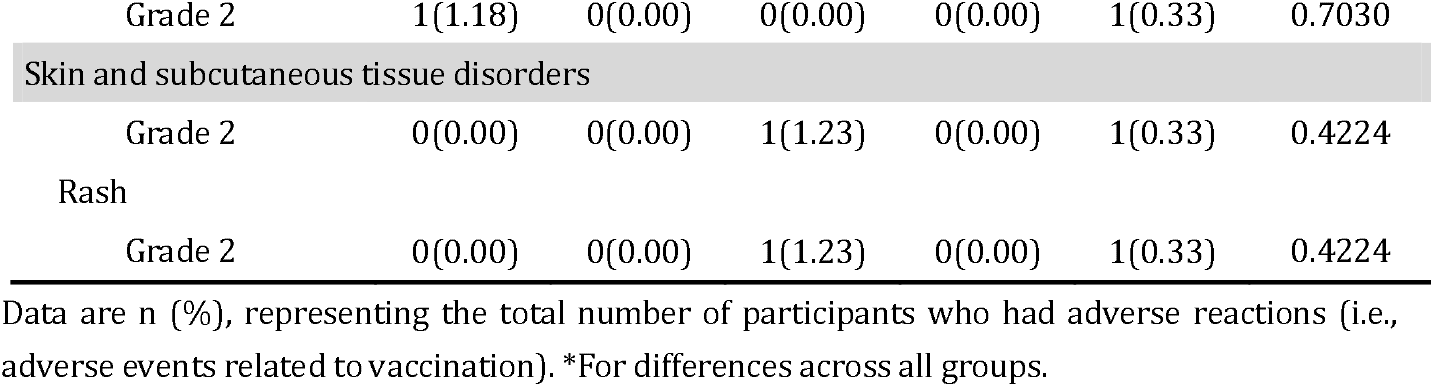
Incidence of adverse reactions within 28 days after the third dose in the phase 2 trial.

| **Adverse Reactions**<br>**(System organ class,**<br>**preferred term)** | **1.5 µg**<br>**group**<br>**(N=85)** | **3 µg**<br>**group**<br>**(N=90)** | **6 µg**<br>**group**<br>**(N=81)** | **Placebo**<br>**group**<br>**(N=47)** | **Total**<br>**(N=303)** | **p value\*** |
| --- | --- | --- | --- | --- | --- | --- |
| **Any adverse reaction** |  |  |  |  |  |  |
| Grade 1 | 3(3.53) | 3(3.33) | 3(3.70) | 2(4.26) | 11(3.63) | 1.0000 |
| Grade 2 | 1(1.18) | 2(2.22) | 2(2.47) | 1(2.13) | 6(1.98) | 0.9468 |
| **Systemic diseases and injection site adverse reactions** |  |  |  |  |  |  |
| Grade 1 | 1(1.18) | 2(2.22) | 2(2.47) | 1(2.13) | 6(1.98) | 0.9468 |
| Grade 2 | 0(0.00) | 2(2.22) | 0(0.00) | 1(2.13) | 3(0.99) | 0.2402 |
| Injection site pain |  |  |  |  |  |  |
| Grade 1 | 1(1.18) | 2(2.22) | 2(2.47) | 1(2.13) | 6(1.98) | 0.9468 |
| Fatigue |  |  |  |  |  |  |
| Grade 2 | 0(0.00) | 1(1.11) | 0(0.00) | 1(2.13) | 2(0.66) | 0.5230 |
| Injection site erythema |  |  |  |  |  |  |
| Grade 2 | 0(0.00) | 1(1.11) | 0(0.00) | 0(0.00) | 1(0.33) | 1.0000 |
| **Respiratory, thoracic and mediastinal disorders** |  |  |  |  |  |  |
| Grade 1 | 1(1.18) | 0(0.00) | 1(1.23) | 1(2.13) | 3(0.99) | 0.5672 |
| Grade 2 | 0(0.00) | 0(0.00) | 1(1.23) | 0(0.00) | 1(0.33) | 0.4224 |
| Cough |  |  |  |  |  |  |
| Grade 1 | 1(1.18) | 0(0.00) | 0(0.00) | 1(2.13) | 2(0.66) | 0.3430 |
| Grade 2 | 0(0.00) | 0(0.00) | 1(1.23) | 0(0.00) | 1(0.33) | 0.4224 |
| Rhinorrhea |  |  |  |  |  |  |
| Grade 1 | 0(0.00) | 0(0.00) | 1(1.23) | 0(0.00) | 1(0.33) | 0.4224 |
| **Nervous system disorders** |  |  |  |  |  |  |
| Grade 1 | 1(1.18) | 1(1.11) | 1(1.23) | 0(0.00) | 3(0.99) | 1.0000 |
| Dizziness |  |  |  |  |  |  |
| Grade 1 | 1(1.18) | 1(1.11) | 0(0.00) | 0(0.00) | 2(0.66) | 1.0000 |
| Headache |  |  |  |  |  |  |
| Grade 1 | 0(0.00) | 0(0.00) | 1(1.23) | 0(0.00) | 1(0.33) | 0.4224 |
| **Gastrointestinal disorders** |  |  |  |  |  |  |
| Grade 1 | 1(1.18) | 1(1.11) | 0(0.00) | 0(0.00) | 2(0.66) | 1.0000 |
| Nausea |  |  |  |  |  |  |
| Grade 1 | 1(1.18) | 1(1.11) | 0(0.00) | 0(0.00) | 2(0.66) | 1.0000 |
| **Musculoskeletal and connective tissue disorders** |  |  |  |  |  |  |
| Grade 2 | 1(1.18) | 0(0.00) | 0(0.00) | 0(0.00) | 1(0.33) | 0.7030 |
| Muscle pain |  |  |  |  |  |  |
| Grade 2 | 1(1.18) | 0(0.00) | 0(0.00) | 0(0.00) | 1(0.33) | 0.7030 |
| Skin and subcutaneous tissue disorders |  |  |  |  |  |  |
| Grade 2 | 0(0.00) | 0(0.00) | 1(1.23) | 0(0.00) | 1(0.33) | 0.4224 |
| Rash |  |  |  |  |  |  |
| Grade 2 | 0(0.00) | 0(0.00) | 1(1.23) | 0(0.00) | 1(0.33) | 0.4224 |

Data are n (%), representing the total number of participants who had adverse reactions (i.e., adverse events related to vaccination). \*For differences across all groups.

**Figure 1.**
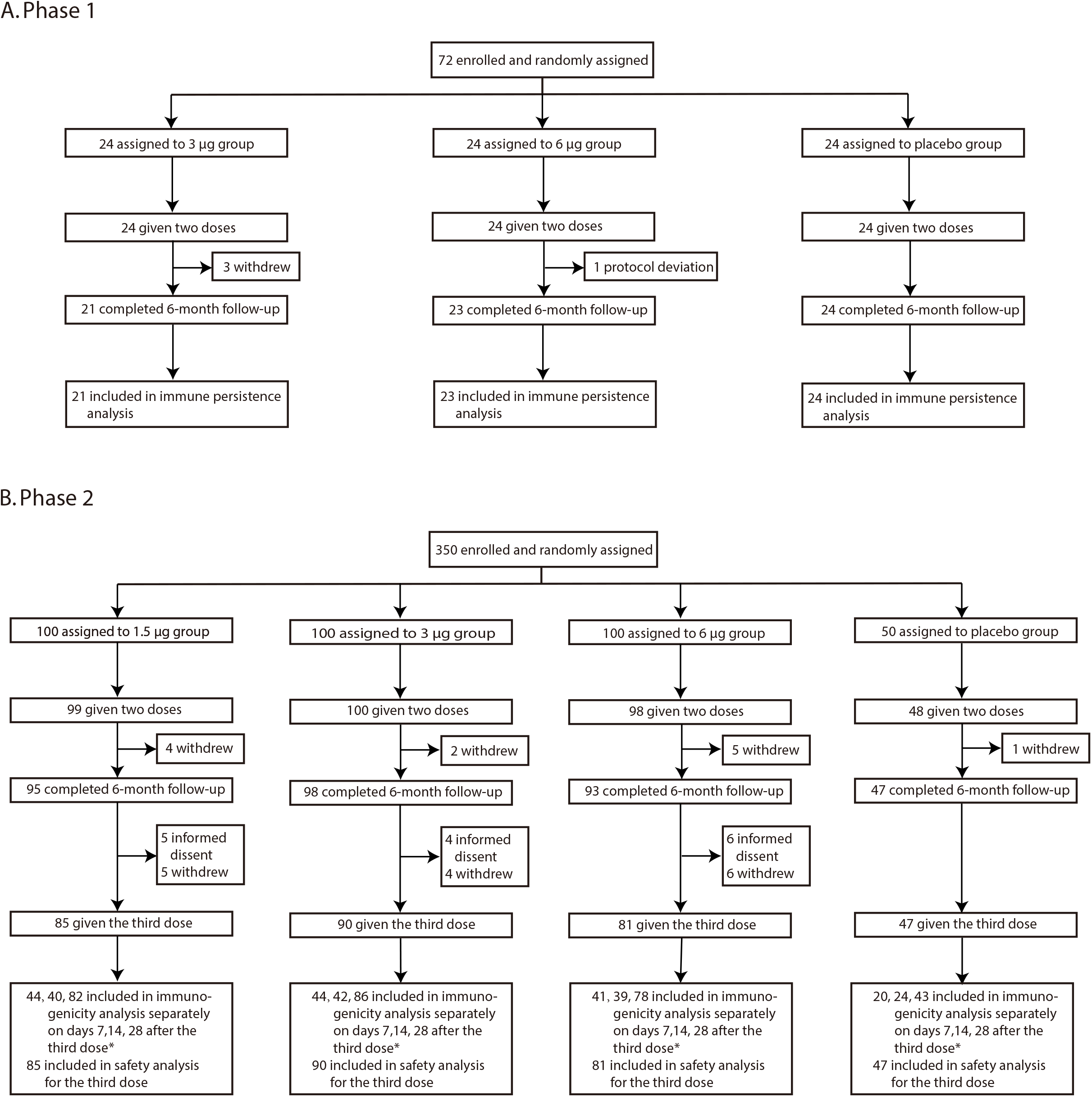
Study Profile. *Participants received the third dose in phase 2 trial; blood samples were taken on day 7 or day 14 (1:1) after the third dose according to randomization code. Blood specimens were obtained on all subjects on day 28 after the third dose.

### Immunogenicity persistence

At baseline, with an exception of 1 of 100 (1.00%) participants in the 6 μg group in the clinical trial phase 2, no participants had detectable neutralizing antibodies. In the placebo group, 1 of 24 (4.17%) participants had detectable neutralizing antibodies in phase,1 and 1 of 47 (2.13%) participants had neutralizing antibodies above seropositive cutoff (titre: 12) in phase 2 at six months after the second dose; while 1 of 43 (2.33%) participants had detectable neutralizing antibodies in phase 2 on day 28 after the third dose (Table S1-S2).

Neutralizing antibody titres declined to below the seropositive cutoff 6 months after two priming doses, and insignificant differences in GMT were observed between vaccine groups in phase 2. GMTs were 3.1 [95%CI 2.7-3.6], 3.4 [95%CI 2.9-4.1], and 4.1 [95%CI 3.3-5.1] separately in the 1.5 μg, 3 μg, and 6 μg vaccine groups, corresponding to seropositivity decreases from over 90% on day 28 after the second dose to 11.76% [95%CI 5.79%-20.57%], 17.78% [95%CI 10.52%-27.26%], and 21.52% [95%CI 13.06%-32.20%] (Figure 2, Table S1) at six months. Patterns were consistent in all phase vaccine 1 groups (Table S2).

**Figure 2.**
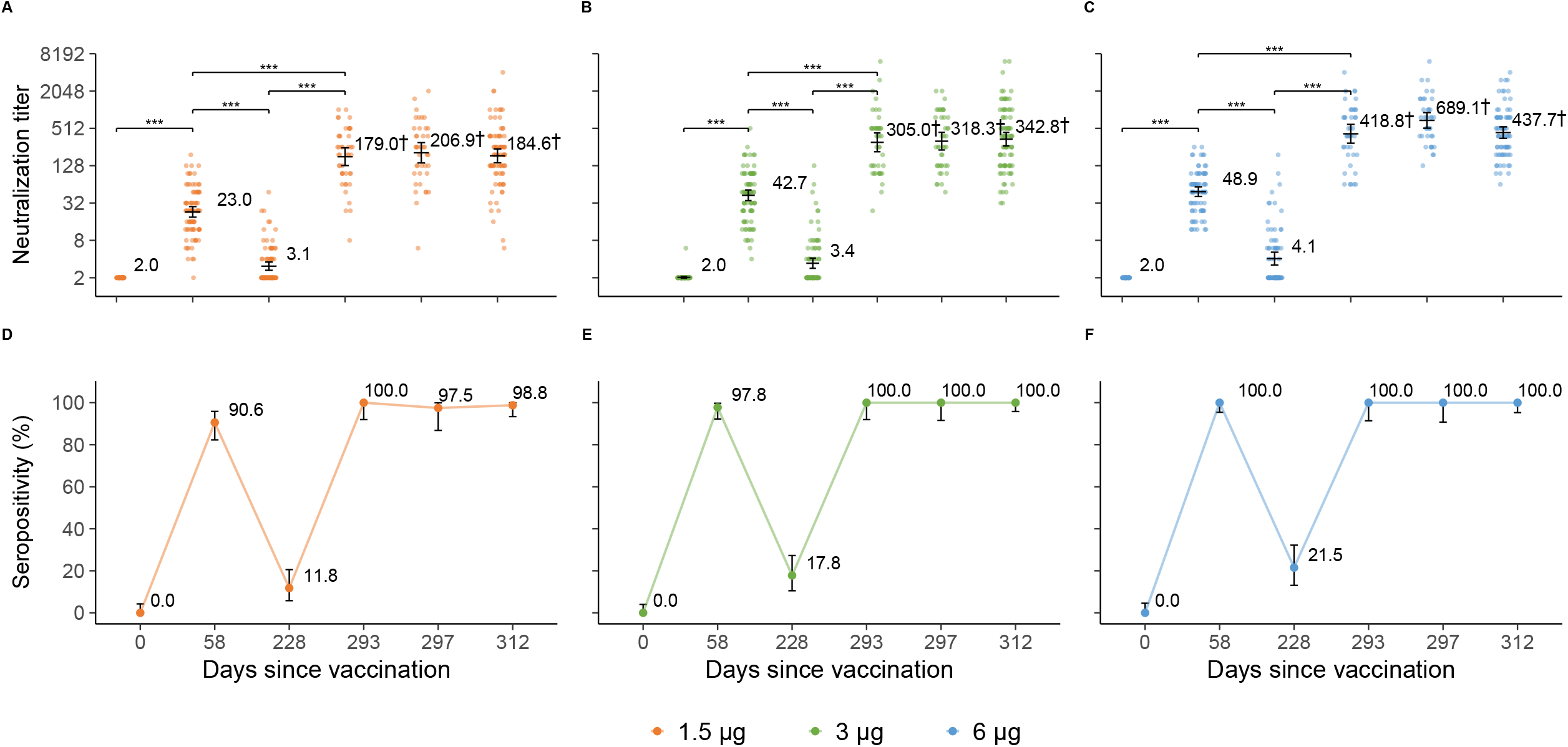
Level of neutralizing antibodies to live SARS-CoV-2 in the phase 2 trial. A: 1.5 μg group; B: 3 μg group; C: 6 μg group. Data are represented as reciprocal neutralizing antibody titres regarding the time after the first dose in the per-protocol population. Numbers above the bars show the Geometric Mean Titre (GMT), and the error bars indicate 95% CIs. Statistical differences were assessed by t-tests on log-transformed data. *p<0.05, **p<0.005, ***p<0.0005, ****p<0.0001. †No significant differences in neutralization titres were observed on days 293, 297 and 312 in the same vaccine group.

### Immunogenicity of the third dose

A third dose given 8 months after the second dose significantly increased neutralizing antibody levels on day 7 post the third dose in all vaccine groups. In the 3 μg group (the licensed formulation), GMTs increased from 3.4 [95%CI 2.9-4.1) six months after the second dose to 305.0 [95%CI 215.3-432.0] on day 7 post the third dose (p<0.001). The GMTs continued to increase on day 14 and on 28 after the third dose, but the differences between day 14/28 and day 7 after the third dose were not statically significant. The GMTs on day 7 after the third dose were approximately 7-fold greater than that on day 28 after the second dose (Figure 2, Table S1). All subjects in the 3 μg group were seropositive after their third doses. No differences in neutralization antibodies level between age groups (60-64 years, 65-69 years, 70 years and older) were observed in age-specific analyses (Figure S1).

Similar patterns of GMTs after third doses were observed for the 1.5 μg group and the 6 μg group. Compared to the 3 μg group on day 28 after the third dose, GMTs in the 6 μg group (437.7 [95%CI 353.8-541.6]) were consistent in direction, but significantly higher than that observed in the 1.5 μg group (184.6 [95%CI 142.8-238.5]). On days 7, 14, and 28 after the third dose, the mean seropositivity rates in the 1.5 μg group ranged from 97.5%-100%, and were 100% in the 3μg and 6 μg groups (Figure 2, Table S1).

### Reactogenicity and safety

Severity grades of solicited local and systemic adverse reactions reported within 28 days after the third dose were grade 1 or grade 2 in all vaccination groups in the study part 2. Incidences of adverse reactions after the third dose were 4.71% (4/85), 5.56% (5/90), 6.17% (5/81), and 4.26% (2/47) for the 1.5 μg, 3 μg, 6 μg, and placebo groups, respectively, with no significant difference between groups (p<0.05) (Table 3, Table S2). The most commonly reported reaction was injection-site pain (Table 3).

In addition of serious adverse events reported in the previous period^12^, a total of 17 serious adverse events occurred in 15 participants until 6 months after the second dose in phase 1/2 trials, including two deaths (one death from cholangiocarcinoma occurred in the 1.5 μg group, one death from hypertensive heart disease occurred in the 6 μg group). From initiation of the third dose until 28 days later in the phase 2 trial, 5 serious adverse events among 4 participants were reported. Non-fatal serious adverse events are detailed in Table S3-S4. None of the serious adverse events were considered by the investigators to be related to vaccination, and no prespecified trial-halting rules were met. Safety monitoring will continue for 1 year after administration of the third vaccine dose in the phase 2 trial.

## Discussion

To our knowledge, this is the first paper to report the duration of the immune response following two-dose CoronaVac schedules and the immunogenicity of a booster dose among adults 60 years or older. We found that neutralizing antibody titres dropped below a seropositive cutoff of 8 by 6 months after two-dose, 28-day interval CoronaVac scheduled vaccination. A third dose, given 8 months after the second dose rapidly and markedly increased neutralizing antibody titres. In the study group that used the licensed dose amount (3 μg), GMTs increased to 305 on the seventh day after vaccination - approximately 7-fold greater than on day 28 after the second dose. We believe that the rapid and strong response is evidence of induction of immune memory by the first two doses of CoronaVac. We found no safety concerns with a booster dose in these older adults, and the reactogenicity of the vaccine was indistinguishable from the reactogenicity of an aluminum hydroxide placebo.

We found that neutralizing antibodies titres dropped below the seropositive cutoff of 8 in over 70% of participants (nearly 90% in the 1.5 μg group) 6 months after primary vaccination. However, we were unable to determine the timing of a change from seropositive to seronegative since we collected no blood specimens between 28 days and 6 months after the second dose. Knowledge of the kinetics of neutralizing antibodies after primary vaccination is useful for determining the optimal timing of a booster dose. Modeling waning of neutralizing antibodies titres, potentially with the use of pooled data from phase 1/2 trials in both older adults and younger adults,^9^ could shed light on booster dose timing. However, the lack of an agreed-upon correlation of protection challenges definitive modelling.

Given the current state of knowledge, it is not possible to know whether there is some degree of protection against COVID-19 after neutralizing antibodies decay to below the seropositive cutoff. In addition to adaptive immune response, innate immune response plays an important role in protection against COVID-19. It was shown that a second dose of the Pfizer-BioNTech vaccine, BNT162b2, stimulates a strikingly enhanced innate immune response^14^. Khoury and colleagues^15^ found that the neutralization level required for protection from severe infection was significantly lower than that for detectable infection. In other words, protection from severe infection was predicted to persist longer than protection against mild infection.

After a booster dose, GMTs in the 3 μg group were identical to the 6 μg group but higher than the 1.5 μg group. The 3 μg formation has been approved for emergency use in children 3-17 years of age, and for conditional use in individuals 18 years of age or older in China^16^; it is also the formulation that has been included in the World Health Organization’s (WHO) emergency use listing^17^. Our study supported it as a booster dose to older adults.

During the first three months of mass vaccination in Chile, a two-dose schedule of 3 μg CoronaVac showed good effectiveness against COVID-19, i.e., having a vaccine effectiveness of 66.6% against symptomatic infection, 85.3% against hospitalization, 89.2% against ICU admission, and 86.5% against death among a population aged 60 years or older, with no significant differences in effectiveness between elderly and younger adults^18^.

In our micro cytopathogenic effect assay, we conducted back titrations to confirm that the amount of virus used in each batch of laboratory tests was within the range of 32-320 TCID50/50 μl. Each batch of laboratory tests was performed with specimens collected in phase 1/2 trials separately in adults aged 60 years or above and in adults younger than 60 years^9^. All tests were done by the National Institute for Food and Drug Control, the laboratory arm of China’s vaccine regulatory authority^12^. The back titration titre was 55 for the elderly adult group, and 247 for the younger adult group, both in an acceptable range. However, the lower back titration titre indicates a lower amount of virus in the elderly adult group, which may lead to higher neutralizing antibody titres. Accordingly, the higher GMT from a booster dose of CoronaVac in the elderly adults than what we observed in adults younger than 60 years of age^9^ does not necessarily provide evidence of better immunogenicity of a booster dose in elderly adults than younger adults; the difference should be interpreted with caution.

The global shortage of COVID-19 vaccine requires wise policy recommendations on how best to use available doses to achieve the greatest public health benefit. Last December, the Joint Committee on Vaccination and Immunisation (JCVI) in the United Kingdom, based on characteristics of their licensed vaccines and epidemic situation, recommended that as many people on the JCVI priority list as possible should be offered a first vaccine dose as an initial priority, even though vaccine efficacy is better after a second dose is administered^19^. In contrast, when using CoronaVac, we suggest ensuring that older adults complete the current two-dose schedule, as this would maximize the impact of the vaccine programme in its primary aim of reducing mortality and hospitalizations and protecting medical institutions. Timing of a booster dose should account for the local epidemic situation, risk of infection, vaccine supply, and other relevant factors.

The incidence of adverse reactions after the third dose was consistently low (around 5%) across all vaccine groups and the placebo group, indicating that a third dose was well-tolerated. Although no major safety concern was identified during our trial, adverse reactions are important to monitor in large-scale clinical trials and during post-marketing periods.

Our study has limitations. First, we only reported immune response data for healthy older adults, and did not include individuals who have underlying medical conditions. The populations are more likely to have co-morbidities placing them at higher risk of severe clinical outcomes. Second, we had only three participants 80 years of age or older, making it not possible to determine immunogenicity results for this age group. Two doses of BNT162b2 induced smaller neutralizing responses in vaccinees over 80 years of age than younger groups,^20,21^ but our study cannot provide evidence one way or the other. Third, high neutralizing titres are important for protection against novel circulating SARS-CoV-2 variants conferring immune escape^22^. We did not perform neutralization testing in vitro against variants to determine the neutralization ability of the vaccine to emerging variants of concern. Fourth, a long-term follow-up of our phase 2 trial will be needed to determine a satisfactory duration of immunity induced by the booster dose.

## Conclusion

Our study found that a two-dose schedule using the licensed dose amount of CoronaVac effectively primes the immune system in adults aged 60 years or older. Although neutralizing antibody titres decreased to low levels 6 months after the second dose, we showed that a third dose given eight months after the second dose rapidly induces a potent immune response. Our results - coupled with additional evidence of vaccine efficacy/effectiveness, epidemic situation, risk of infection, and vaccine supply - can help policymakers optimize the timing of a booster dose of CoronaVac for the greatest public health impact.

## Data Availability

All the data referred to in the manuscript will be provided with reasonable request after the trial is completed.

## Declarations

### Competing interests

H.Y. has received research funding from Sanofi Pasteur, GlaxoSmithKline, Yichang HEC Changjiang Pharmaceutical Company, and Shanghai Roche Pharmaceutical Company. None of those research funding is related to development of COVID-19 vaccines. L.W. and Y.W. were the employees of Sinovac Biotech Ltd., L.W. was an employee of Sinovac Life Sciences Co., Ltd. All other authors report no competing interests.

### Funding

The study was supported by grants from National Key Research and Development Program (2020YFC0849600), Beijing Science and Technology Program (Z201100005420023), and National Science Fund for Distinguished Young Scholars (81525023).

## Acknowledgements

We thanked Dr. Lance Rodewald from Chinese Center for Disease Control and Prevention for his kind comments on this paper.

## Notes

### Clinical Trial

ClinicalTrials.gov (NCT04383574)

### Author Declarations

Written informed consent was obtained from each participant before the third dose in the second part of the study. The clinical trial protocol and informed consent form were approved by the Ethics Committee of Hebei CDC (IRB2020-006).

