## Supplementary for "A booster dose is immunogenic and will be needed for older adults who have completed two doses vaccination with CoronaVac: a randomised, double-blind, placebo-controlled, phase 1/2 clinical trial"

Supplemental Methods

Appendix 1. Exclusion criteria for the administration of the third dose

- **Continuing vaccination is prohibited, and other research steps may continue according to researchers’ judgement of:**

(1) A COVID-19 vaccine other than the experimental vaccine was used during the study period;

(2) Any serious vaccine-caused adverse reactions are observed in the subject;

(3) Severe anaphylaxis or hypersensitivity after vaccination (including urticaria / rash within 30 minutes after vaccination);

(4) Any confirmed or suspected autoimmune or immunodeficiency disease, including human immunodeficiency virus (HIV) infection;

- **Vaccination can be postponed within the time window specified in the program if:**

(5) Acute or new chronic diseases happen after vaccination;

(6) Other reactions (including severe pain, severe swelling, severe activity limitation, persistent high fever, severe headache or other systemic or local reactions) occur, as judged by the researcher;

- **Vaccination can be postponed within the time window specified in the protocol if:**

(7) At the time of vaccination, the participant was suffering from acute disease (acute disease refers to moderate or severe disease with or without fever);

(8) The axillary temperature was higher than 37.0℃;

(9) Participants were vaccinated with a non-COVID-19 subunit vaccine or inactivated vaccine within 7 days or attenuated vaccine within 14 days;

(10) According to the judgment of the researcher, the participant had any other factors not suitable for vaccination.

Appendix 2: Trial profile

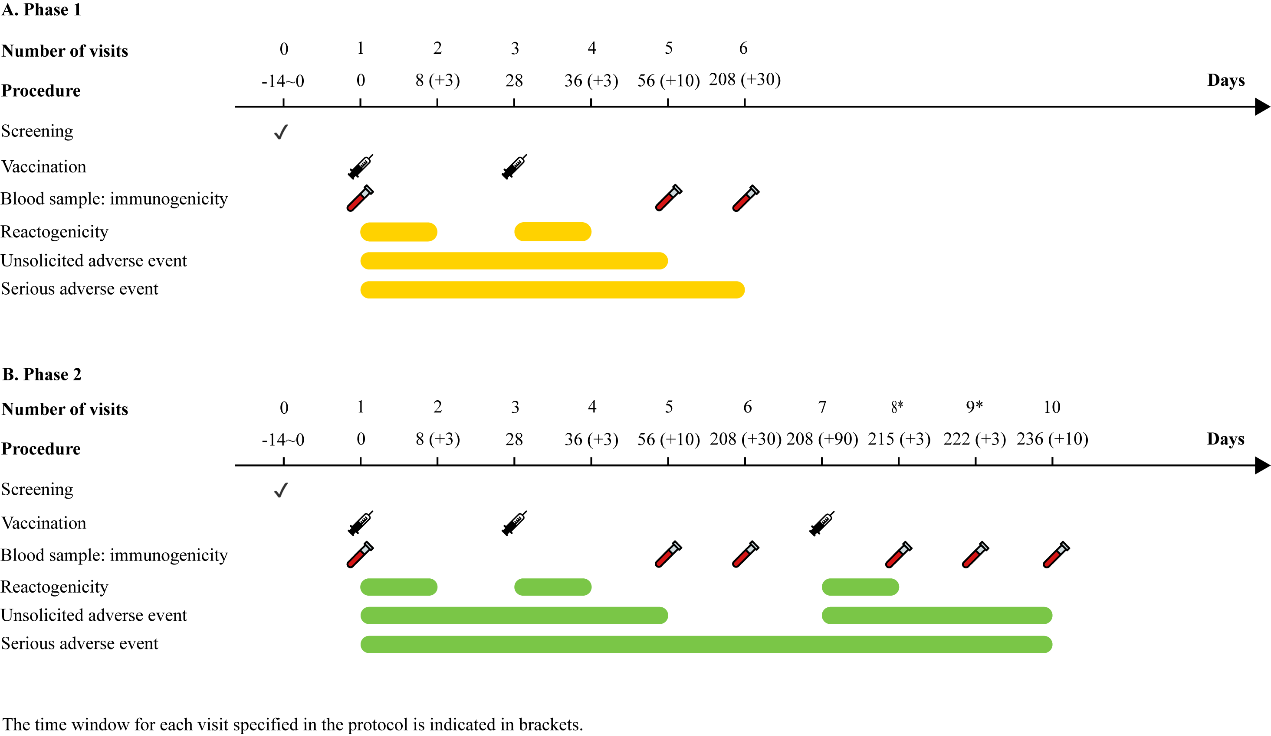

*The first 175 participants had blood samples taken on day 215, and the next 175 participants had blood samples taken on day 222, in accordance with their randomization codes.

Appendix 3: Descriptions of data sets for immunogenicity analysis

**Phase 1**

**Immune Persistence Set (IPS-6)**

IPS-6 included subjects who completed the two-dose schedule immunization and completed blood sampling for immunogenicity evaluation at 6 months after the second dose.

**Phase 2**

**Full Analysis Set for the Third Dose (bFAS)**

bFAS included subjects who received the third dose of CoronaVac or placebo, completed at least one blood sampling before and after vaccination for immunogenicity evaluation following the principle of intention to treatment (ITT). Subjects with incorrect vaccination were evaluated for immunogenicity in original randomized groups according to ITT principles.

**Per Protocol Set for the Third Dose (bPPS)**

bPPS was a subset of bFAS and included all subjects who met enrolment criteria and received two-dose primary immunization and the third dose after 6 months within the time window according to the protocol requirements. Blood samples of subjects were collected before the booster 28 days after the third dose, and evaluated for immunogenicity effectively. Exclusion criteria for bPPS was as follows:

- subject violated protocol;
- subjects given wrong interventional product;
- the injection of booster dose or the blood sampling 28 days after the third dose fell outside the time window;
- subjects using vaccines or drugs prohibited in protocol, including
  - other research or unregistered products (drugs or vaccines) in addition of investigational product,
  - long-term use (more than 14 days) of immunosuppressants or other immunomodulatory drugs (inhaled or topical steroids are allowed),
  - immunoglobulins and / or blood preparations.
- subjects with newly diagnosed autoimmune diseases or immune disorders, including human immunodeficiency virus (HIV) infection;
- other factors affecting the immunogenicity evaluation of the vaccine.

**Per Protocol Set 1 for the Third Dose (PPS1)**

PPS1 was a subset of bFAS and included all subjects who met enrolment criteria and received two-dose primary series doses and the third dose after 6 months within the time window according to the protocol requirements. Blood samples of subjects were collected before the booster 7 days after the third dose, and evaluated for immunogenicity. Exclusion criteria for PPS1 was as follows:

- subject violated protocol;
- subject given wrong interventional product;
- booster dose administration or blood sampling 7 days after the third dose fell outside the time window;
- subjects used vaccines or drugs prohibited in protocol, including
  - other research or unregistered products (drugs or vaccines) in addition of investigational product,
  - long-term use (more than 14 days) of immunosuppressants or other immunomodulatory drugs (inhaled or topical steroids are allowed),
  - immunoglobulins and / or blood preparations.
- subjects with newly diagnosed autoimmune diseases or immune system diseases, including human immunodeficiency virus (HIV) infection;
- other factors affecting the immunogenicity evaluation.

**Per Protocol Set 2 for the Third Dose (PPS2)**

PPS2 was a subset of bFAS and included all subjects who met enrolment criteria and received two-dose primary immunization and the third dose after 6 months within the time window according to the protocol requirements. Blood samples of subjects were collected before the booster 14 days after the third dose, and evaluated for immunogenicity effectively. Exclusion criteria for PPS2 was as follows:

- subject violated protocol;
- subject given wrong interventional product;
- booster dose administration or blood sampling 14 days after the third dose fell outside the time window;
- subjects used vaccines or drugs prohibited in protocol, including
  - other research or unregistered products (drugs or vaccines) in addition of investigational product,
  - long-term use (more than 14 days) of immunosuppressants or other immunomodulatory drugs (inhaled or topical steroids are allowed),
  - immunoglobulins and / or blood preparations.
- subjects with newly diagnosed autoimmune diseases of immune system diseases, including human immunodeficiency virus (HIV) infection;
- other factors affecting the immunogenicity evaluation.

Supplemental Results of immunogenicity

Table S1. Level of neutralizing antibodies to live SARS-CoV-2 in the phase 2 trial (per-protocol analysis)

| **Days from**  **vaccination** | **Indicators** | **1.5 μg group** | **3 μg** **group** | **6 μg** **group** | **placebo** **group** | **P value** | | | |
| --- | --- | --- | --- | --- | --- | --- | --- | --- | --- |
|  |  |  |  |  |  | **All** | **1.5 μg vs 3 μg** | **1.5 μg vs 6μg** | **3 μg vs 6 μg** |
| **0 (0-0)** | **N** | **85** | **90** | **79** | **47** |  |  |  |  |
| **(Baseline)** | Seropositivity, n (%) | 0 (0) | 0 (0) | 0 (0) | 0 (0) | 1.0000 | **—** | **—** | **—** |
| (FAS) | (95%CI) | (0, 4.25) | (0, 4.02) | (0, 4.56) | (0, 7.55) |  |  |  |  |
|  | GMT* | 2.0 | 2.0 | 2.0 | 2.0 | 0.506 | **—** | **—** | **—** |
|  | (95%CI) | (2.0, 2.0) | (2.0, 2.1) | (2.0, 2.0) | (2.0, 2.0) |  |  |  |  |
| **58 (56-72)** | **N** | **85** | **90** | **79** | **47** |  |  |  |  |
| (V2+28) | Seropositivity, n (%) | 77 (90.59) | 88 (97.78) | 79 (100.00) | 0 (0) | <0.0001 | 0.8081 | 0.7414 | 1.0000 |
| (FAS) | (95%CI) | (82.29, 95.85) | (92.20, 99.73) | (95.44, 100.00) | (0, 7.55) |  |  |  |  |
|  | GMT^*^ | 23.0 | 42.7 | 48.9 | 2.1 | <0.0001 | <0.0001 | <0.0001 | 1.0000 |
|  | (95%CI) | (18.9, 28.0) | (35.0, 52.0) | (40.9, 58.4) | (2.0, 2.1) |  |  |  |  |
| **228 (219-235)** | **N** | **85** | **90** | **79** | **47** |  |  |  |  |
| (V2+180) | Seropositivity, n (%) | 10 (11.76) | 16 (17.78) | 17 (21.52) | 1 (2.13) | 0.0289 |  |  |  |
| (FAS) | (95%CI) | (5.79, 20.57) | (10.52, 27.26) | (13.06, 32.20) | (0.05, 11.29) |  |  |  |  |
|  | GMT* | 3.1 | 3.4 | 4.1 | 2.1 | 0.0002 | 1.0000 | 0.1966 | 1.0000 |
|  | (95%CI) | (2.6, 3.6) | (2.8, 4.1) | (3.2, 5.2) | (1.9, 2.2) |  |  |  |  |
| **293 (291-294)** | **N** | **44** | **44** | **41** | **20** |  |  |  |  |
| (V3+7) | Seropositivity, n (%) | 44 (100.00) | 44 (100.00) | 100 (100.00) | 0 (0) | <0.0001 | 1.0000 | 1.0000 | 1.0000 |
| (PPS1) | (95%CI) | (91.96, 100.00) | (91.96, 100.00) | (91.40, 100.00) | (0.00, 16.84) |  |  |  |  |
|  | GMT* | 179.0 | 305.0 | 418.8 | 2.0 | <0.0001 | 0.1024 | 0.0014 | 0.9636 |
|  | (95%CI) | (128.7, 248.9) | (215.3, 432.0) | (295.6, 593.3) | (2.0, 2.0) |  |  |  |  |
| **297 (297-299)** | **N** | **40** | **42** | **38** | **24** |  |  |  |  |
| (V3+14) | Seropositivity, n (%) | 39 (97.50) | 42 (100.00) | 38 (100.00) | 0 (0) | <0.0001 | 1.0000 | 1.0000 | 1.0000 |
| (PPS2) | (95%CI) | (86.84, 99.94) | (91.59, 100.00) | (90.75, 100.00) | (0.00, 14.25) |  |  |  |  |
|  | GMT* | 206.9 | 318.3 | 689.1 | 2.0 | <0.0001 | 0.2588 | <0.0001 | 0.0025 |
|  | (95%CI) | (142.0, 301.4) | (229.6, 441.2) | (515.9, 920.4) | (2.0, 2.0) |  |  |  |  |
| **312 (311-314)** | **N** | **82** | **86** | **76** | **43** |  |  |  |  |
| (V3+28) | Seropositivity, n (%) | 81 (98.78) | 86 (100.00) | 76 (100.00) | 0 (0) | <0.0001 | 1.0000 | 1.0000 | 1.0000 |
| (PPS) | (95%CI) | (93.39, 99.97) | (95.80, 100.00) | (95.26, 100.00) | (0.00, 8.22) |  |  |  |  |
|  | GMT* | 184.6 | 342.8 | 437.7 | 2.0 | <0.0001 | 0.0006 | <0.0001 | 0.77043 |
|  | (95%CI) | (142.8, 238.5) | (266.4, 441.1) | (353.8, 541.6) | (2.0, 2.1) |  |  |  |  |

* Statistical differences were assessed by t-test on log-transformed data.

Days from vaccination were shown as median (range) days between each visit and the first dose.

Table S2. Level of neutralizing antibodies to live SARS-CoV-2 in the phase 1 trial (IPS-6)

| **Days from**  **vaccination** | **Indicators** | **3 μg** **group** | **6 μg** **group** | **placebo** **group** | **P value** | |
| --- | --- | --- | --- | --- | --- | --- |
|  |  | **N=21** | **N=23** | **N=24** | **All** | **3 μg vs 6 μg** |
| **0 (0-0)** | Seropositivity, n (%) | 0 (0) | 0 (0) | 0 (0) | 1.0000 |  |
| (Baseline) | (95%CI) | (0.00, 16.11) | (0.00, 14.82) | (0.00, 14.25) |  |  |
|  | GMT* | 2.0 | 2.0 | 2.0 | 1.0000 |  |
|  | (95%CI) | (2.0, 2.0) | (2.0, 2.0) | (2.0, 2.0) |  |  |
| **28 (28-28)** | Seropositivity, n (%) | 10 (47.6) | 14 (60.87) | 0 (0.00) | 0.0007 | 0.7994 |
| (V1+28) | (95%CI) | (25.71, 70.22) | (38.54, 80.29) | (0.00, 14.25) |  |  |
|  | GMT* | 6.4 | 8.7 | 2.0 | ＜0.0001 | 0.2594 |
|  | (95%CI) | (4.1, 9.9) | (6.1, 12.5) | (2.0, 2.0) |  |  |
| **58 (56-72)** | Seropositivity, n (%) | 21 (100.00) | 22 (95.65) | 0 (0.00) | ＜0.0001 | 1.0000 |
| V2+28 | (95%CI) | (83.89, 100.00) | (78.05, 99.89) | (0.00, 14.25) |  |  |
|  | GMT* | 54.2 | 64.4 | 2.0 | ＜0.0001 | 0.5500 |
|  | (95%CI) | (36.2, 81.1) | (41.5 99.7) | (2.0, 2.0) |  |  |
| **222 (210-243)** | Seropositivity, n (%) | 6 (28.57) | 3 (13.04) | 1 (4.17) | 0.1424 | 0.4672 |
| (V2+180) | (95%CI) | (11.28,52.18) | (2.78, 33.59) | (0.11,21.12) |  |  |
|  | GMT* | 5.0 | 3.8 | 2.2 | 0.0004 | 0.2602 |
|  | (95%CI) | (3.4,7.3) | (2.9,5.0) | (1.9,2.7) |  |  |

IPS-6 represents the immune persistence set. *Statistical differences were assessed by t-test on log-transformed data.

Days from vaccination were shown as median (range) days between each visit and the first dose.

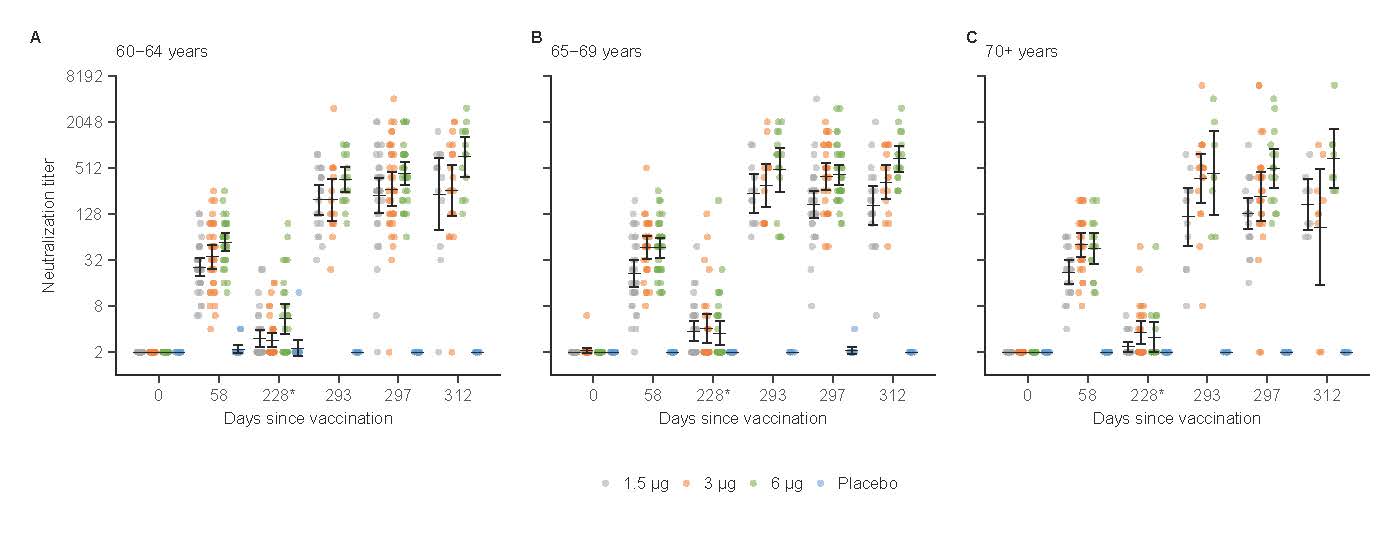

Figure S1. Neutralising antibody responses to live SARS-CoV-2 by age groups in the phase 2 trial

Supplemental Results of safety

Table S3. Overview of adverse reactions reported within 28 days after the third dose in the phase 2 trial

| Classification | 1.5 μg group  (N=85) | | 3 μg group  (N=90) | | 6 μg group  (N=81) | | Placebo group  (N=47) | | Total  (N=303) | | *P* value* |
| --- | --- | --- | --- | --- | --- | --- | --- | --- | --- | --- | --- |
|  | No. of  events | No. of  subjects (%) | No. of  events | No. of  subjects (%) | No. of  events | No. of  subjects (%) | No. of  events | No. of  subjects (%) | No. of  events | No. of  subjects (%) |  |
| Total | 5 | 4(4.71) | 6 | 5(5.56) | 6 | 5(6.17) | 3 | 2(4.26) | 20 | 16(5.28) | 0.9839 |
| Local reactions | 1 | 1(1.18) | 3 | 3(3.33) | 2 | 2(2.47) | 1 | 1(2.13) | 7 | 7(2.31) | 0.9130 |
| Systemic reactions | 4 | 3(3.53) | 3 | 2(2.22) | 4 | 3(3.70) | 2 | 2(4.26) | 13 | 10(3.30) | 0.8702 |
| Solicited events | 4 | 4(4.71) | 5 | 5(5.56) | 5 | 5(6.17) | 3 | 2(4.26) | 17 | 16(5.28) | 0.9839 |
| Unsolicited events | 1 | 1(1.18) | 1 | 1(1.11) | 1 | 1(1.23) | 0 | 0(0.00) | 3 | 3(0.99) | 1.0000 |
| Within 30 minutes | 1 | 1(1.18) | 1 | 1(1.11) | 0 | 0(0.00) | 0 | 0(0.00) | 2 | 2(0.66) | 1.0000 |
| Day 0~7 | 5 | 4(4.71) | 6 | 5(5.56) | 6 | 5(6.17) | 3 | 2(4.26) | 20 | 16(5.28) | 0.9839 |

**P* value was calculated with the Fisher’s exact test

Table S4. Serious adverse events reported in the phase 1 trial

| **Adverse Events**  **(System organ class, preferred term)** | 3 μg group (N=24) | | 6 μg group (N=24) | | Placebo group (N=24) | | Total (N=72) | | *P* value* |
| --- | --- | --- | --- | --- | --- | --- | --- | --- | --- |
|  | *No. of*  *events* | *No. of*  *subjects (%)* | *No. of*  *events* | *No. of*  *subjects (%)* | *No. of*  *events* | *No. of*  *subjects (%)* | *No. of*  *events* | *No. of*  *subjects (%)* |  |
| **Total** | 0 | 0(0.00) | 7 | 3(12.5) | 0 | 0(0.00) | 7 | 3(4.17) | 0.1018 |
| **Heart diseases** |  |  |  |  |  |  |  |  |  |
| coronary artery disease | 0 | 0(0.00) | 1 | 1(4.17) | 0 | 0(0.00) | 1 | 1(1.39) | 1.0000 |
| coronary sclerosis | 0 | 0(0.00) | 1 | 1(4.17) | 0 | 0(0.00) | 1 | 1(1.39) | 1.0000 |
| **Vascular and lymphatic diseases** | | | | | | | | | |
| Hypertension | 0 | 0(0.00) | 2 | 2(8.33) | 0 | 0(0.00) | 2 | 2(2.78) | 0.3239 |
| **Nervous system disorders** |  |  |  |  |  |  |  |  |  |
| Hypoxic ischemic encephalopathy | 0 | 0(0.00) | 1 | 1(4.17) | 0 | 0(0.00) | 1 | 1(1.39) | 1.0000 |
| **Skin and Subcutaneous Tissue Disorders** | | | | | | | | | |
| Henoch-Schoenlein purpura | 0 | 0(0.00) | 1 | 1(4.17) | 0 | 0(0.00) | 1 | 1(1.39) | 1.0000 |
| **Kidneys and urinary system disease** | | | | | | | | | |
| Kidney injury | 0 | 0(0.00) | 1 | 1(4.17) | 0 | 0(0.00) | 1 | 1(1.39) | 1.0000 |
| **P* value was calculated with the Fisher’s exact test. | | | | | | | | | |

Table S5. Serious adverse events reported in the phase 2 trial

| **Adverse Events**  **(System organ class, preferred term)** | 1.5 μg group  (N=100) | | 3 μg group  (N=101) | | 6 μg group  (N=99) | | Placebo group  (N=49) | | Total  (N=349) | |  |
| --- | --- | --- | --- | --- | --- | --- | --- | --- | --- | --- | --- |
|  | *No. of*  *events* | *No. of*  *subjects* | *No. of*  *events* | *No. of*  *subjects* | *No. of*  *events* | *No. of*  *subjects* | *No. of*  *events* | *No. of*  *subjects* | *No. of*  *events* | *No. of*  *subjects* | *P* value* |
| **Total** | 13 | 10(10.00) | 5 | 5(4.95) | 8 | 7(7.07) | 2 | 2(4.08) | 28 | 24(6.88) | 0.4934 |
| **Nervous system disorders** | 2 | 2(2.00) | 2 | 2(1.98) | 1 | 1(1.01) | 0 | 0(0.00) | 5 | 5(1.43) | 1.0000 |
| Cerebral infarction | 1 | 1(1.00) | 1 | 1(0.99) | 0 | 0(0.00) | 0 | 0(0.00) | 2 | 2(0.57) | 1.0000 |
| Hypoxic-Ischemic encephalopathy | 1 | 1(1.00) | 0 | 0(0.00) | 1 | 1(1.01) | 0 | 0(0.00) | 2 | 2(0.57) | 0.6690 |
| Transient ischemic attack | 0 | 0(0.00) | 1 | 1(0.99) | 0 | 0(0.00) | 0 | 0(0.00) | 1 | 1(0.29) | 1.0000 |
| **Neoplasms benign, malignant and unspecified (incl cysts and polyps)** | 4 | 4(4.00) | 0 | 0(0.00) | 0 | 0(0.00) | 0 | 0(0.00) | 4 | 4(1.15) | 0.0221 |
| Lung neoplasm | 1 | 1(1.00) | 0 | 0(0.00) | 0 | 0(0.00) | 0 | 0(0.00) | 1 | 1(0.29) | 0.7106 |
| Lung adenocarcinoma | 1 | 1(1.00) | 0 | 0(0.00) | 0 | 0(0.00) | 0 | 0(0.00) | 1 | 1(0.29) | 0.7106 |
| Liver cancer | 1 | 1(1.00) | 0 | 0(0.00) | 0 | 0(0.00) | 0 | 0(0.00) | 1 | 1(0.29) | 0.7106 |
| Kidney cyst | 1 | 1(1.00) | 0 | 0(0.00) | 0 | 0(0.00) | 0 | 0(0.00) | 1 | 1(0.29) | 0.7106 |
| Gastrointestinal disorders | 1 | 1(1.00) | 1 | 1(0.99) | 1 | 1(1.01) | 1 | 1(2.04) | 4 | 4(1.15) | 0.8363 |
| Duodenal Ulcer | 0 | 0(0.00) | 0 | 0(0.00) | 0 | 0(0.00) | 1 | 1(2.04) | 1 | 1(0.29) | 0.1404 |
| Gastrointestinal perforation | 1 | 1(1.00) | 0 | 0(0.00) | 0 | 0(0.00) | 0 | 0(0.00) | 1 | 1(0.29) | 0.7106 |
| Gastritis | 0 | 0(0.00) | 0 | 0(0.00) | 1 | 1(1.01) | 0 | 0(0.00) | 1 | 1(0.29) | 0.4241 |
| Pancreatitis | 0 | 0(0.00) | 1 | 1(0.99) | 0 | 0(0.00) | 0 | 0(0.00) | 1 | 1(0.29) | 1.0000 |
| **Respiratory, thoracic and mediastinal disorders** | 1 | 1(1.00) | 0 | 0(0.00) | 2 | 2(2.02) | 0 | 0(0.00) | 3 | 3(0.86) | 0.3637 |
| Pulmonary inflammation | 0 | 0(0.00) | 0 | 0(0.00) | 1 | 1(1.01) | 0 | 0(0.00) | 1 | 1(0.29) | 0.4241 |
| Pulmonary emphysema | 1 | 1(1.00) | 0 | 0(0.00) | 0 | 0(0.00) | 0 | 0(0.00) | 1 | 1(0.29) | 0.7106 |
| Chronic obstructive  pulmonary disease | 0 | 0(0.00) | 0 | 0(0.00) | 1 | 1(1.01) | 0 | 0(0.00) | 1 | 1(0.29) | 0.4241 |
| **Cardiac disorders** | 3 | 3(3.00) | 0 | 0(0.00) | 0 | 0(0.00) | 0 | 0(0.00) | 3 | 3(0.86) | 0.0982 |
| Unstable angina pectoris | 2 | 2(2.00) | 0 | 0(0.00) | 0 | 0(0.00) | 0 | 0(0.00) | 2 | 2(0.57) | 0.4228 |
| Atrial fibrillation | 1 | 1(1.00) | 0 | 0(0.00) | 0 | 0(0.00) | 0 | 0(0.00) | 1 | 1(0.29) | 0.7106 |
| **Musculoskeletal and connective tissue disorders** | 0 | 0(0.00) | 0 | 0(0.00) | 2 | 2(2.02) | 0 | 0(0.00) | 2 | 2(0.57) | 0.1791 |
| Osteoarthritis | 0 | 0(0.00) | 0 | 0(0.00) | 1 | 1(1.01) | 0 | 0(0.00) | 1 | 1(0.29) | 0.4241 |
| Lumbar spinal stenosis | 0 | 0(0.00) | 0 | 0(0.00) | 1 | 1(1.01) | 0 | 0(0.00) | 1 | 1(0.29) | 0.4241 |
| **Vascular and lymphatic diseases** | 0 | 0(0.00) | 1 | 1(0.99) | 1 | 1(1.01) | 0 | 0(0.00) | 2 | 2(0.57) | 0.8337 |
| Hypertension | 0 | 0(0.00) | 1 | 1(0.99) | 1 | 1(1.01) | 0 | 0(0.00) | 2 | 2(0.57) | 0.8337 |
| **Hepatobiliary disease** | 0 | 0(0.00) | 0 | 0(0.00) | 1 | 1(1.01) | 0 | 0(0.00) | 1 | 1(0.29) | 0.4241 |
| Acute cholecystitis | 0 | 0(0.00) | 0 | 0(0.00) | 1 | 1(1.01) | 0 | 0(0.00) | 1 | 1(0.29) | 0.4241 |
| **Infection and infectious diseases** | 1 | 1(1.00) | 0 | 0(0.00) | 0 | 0(0.00) | 0 | 0(0.00) | 1 | 1(0.29) | 0.7106 |
| Bronchitis | 1 | 1(1.00) | 0 | 0(0.00) | 0 | 0(0.00) | 0 | 0(0.00) | 1 | 1(0.29) | 0.7106 |
| **Injury, poisoning and operative complications** | 1 | 1(1.00) | 0 | 0(0.00) | 0 | 0(0.00) | 0 | 0(0.00) | 1 | 1(0.29) | 0.7106 |
| Tibial fracture | 1 | 1(1.00) | 0 | 0(0.00) | 0 | 0(0.00) | 0 | 0(0.00) | 1 | 1(0.29) | 0.7106 |
| **Surgical and medical procedures** | 0 | 0(0.00) | 1 | 1(0.99) | 0 | 0(0.00) | 0 | 0(0.00) | 1 | 1(0.29) | 1.0000 |
| Hip arthroplasty | 0 | 0(0.00) | 1 | 1(0.99) | 0 | 0(0.00) | 0 | 0(0.00) | 1 | 1(0.29) | 1.0000 |
| **Reproductive system and breast disease** | 0 | 0(0.00) | 0 | 0(0.00) | 0 | 0(0.00) | 1 | 1(2.04) | 1 | 1(0.29) | 0.1404 |
| Benign prostatic hyperplasia | 0 | 0(0.00) | 0 | 0(0.00) | 0 | 0(0.00) | 1 | 1(2.04) | 1 | 1(0.29) | 0.1404 |

Data are n (%). * For differences across all groups.
